# Excess Mortality during the Covid-19 pandemic: Early evidence from England and Wales

**DOI:** 10.1101/2020.04.14.20065706

**Authors:** Sotiris Vandoros

**Affiliations:** King’s College London and Harvard T.H. Chan School of Public Health

**Keywords:** Covid-19, excess mortality, underreporting, lockdown, spillover effects, collateral damage

## Abstract

**Background:** The Covid-19 pandemic has claimed many lives in the UK and globally. The objective of this paper is to study whether the number of deaths not registered as covid-19-related has increased compared to what would have been expected in the absence of the pandemic. This may be a result of some covid-19 deaths being unreported or spillover effects on other causes of death (or both). Reasons behind this might include covid-19 underreporting, avoiding visits to hospitals or GPs, and the effects of the lockdown.

**Methods:** I used weekly ONS data on the number of deaths in England and Wales that did not officially involve covid-19 over the period 2015-2020. Simply observing trends is not sufficient as spikes in deaths may occasionally occur. I thus followed a differences-in-differences econometric approach to study whether there was a relative increase in deaths not registered as covid-19-related during the pandemic, compared to a control. As an additional approach, an interrupted time series model was also used.

**Results:** Results suggest that there are an additional 968 weekly deaths that officially did not involve covid-19, compared to what would have otherwise been expected. This increase is also confirmed by the interrupted time series analysis.

**Discussion:** The number of deaths not officially involving covid-19 has demonstrated an absolute and relative increase during the pandemic. It is possible that some people are dying from covid-19 without being diagnosed, and that there are excess deaths due to other causes as a result of the pandemic. Analysing the cause of death for any excess non-covid-19 deaths will shed light upon the reasons for the increase in such deaths and will help design appropriate policy responses to save lives.

## 1. Background

Over 4 million Covid-19 cases have been reported globally, leading to 300,000 deaths. In the United Kingdom, the death toll has reached 33,000, while over 220,000 people have been diagnosed with the virus.^1^ The novel coronavirus is directly claiming lives, and it is also possible that some covid-19 patients may have died without being diagnosed. However, this unprecedented situation and the lockdown might also be triggering additional health problems. People with other, unrelated health conditions may be reluctant to visit their GP or a hospital in order to avoid the risk of contracting the virus,^2^ thus remaining undiagnosed or not receiving the medical treatment they might need. Furthermore, to increase capacity for the overstretched NHS, routine operations have been postponed.^3^ The lockdown may also have unintended health effects. Lack of social contact can affect mental health,^4^ and big events or disasters at the national level can have a similar impact.^5^ Staying at home can limit physical activity, which has been associated with obesity^6^ and mental health.^7^ There are also reports of a rise in domestic abuse,^8^ while the current financial and public health situation may also cause additional uncertainty and stress.^9-10^ Apart from the negative effects, there may also be some improvement in certain areas. The lockdown has reduced traffic volume and may thus lead to a decrease in motor vehicle collisions and related deaths. Reduced traffic has also led to lower levels of air pollution, which is associated with mortality.^11^ The lockdown may have also helped reduce crime rates.

The objective of this paper is to study whether and to what extent the number of deaths not registered as covid-19-related have increased compared to what would have been expected in the absence of the virus. This may be a result of some covid-19 deaths being unreported or spillover effects on other causes of death (or both).

## 2. Data and Methods

This study used weekly (provisional) mortality data from England and Wales for years 2019 and 2020, obtained from the Office for National Statistics (ONS).^12^ Data were extracted on 7 April 2020 and updated on 14, 21, 28 April and on 5 and 12 May 2020. Data used in this study are based on the data released on 12 May 2020, and values included in this dataset may be changed in later releases, as is sometimes the case. Data were reported by gender, age group and Region. I used the total number of deaths (regardless of cause) as well as the number of deaths where Covid-19 was mentioned on the death certificate, in order to calculate the number of deaths that were not officially related to Covid-19. Data on Covid-19 deaths are also available by the Department of Health and Social Care,^13^ but the latter exclude those deaths that occurred outside hospital, which is why I preferred the ONS data. According to the ONS, data by gender or age group may be incomplete, so they might not necessarily sum to the total number of deaths.

Studying trends in a variable alone before and after a “treatment” can be misleading as there may be other factors driving any change. For that purpose, a control group can help filter out any other effects. Such a control group will have to remain unaffected by the treatment. The covid-19 pandemic is a major global crisis, and has caused a lockdown throughout the UK, so identifying a control population for the same period seems impossible as it would be highly likely to be contaminated. Instead, I follow an approach similar to that by Metcalfe et al^5^ and Powdthavwee et al^14^ who used trends in the same variable, in earlier years, as a control group. Likewise, I used deaths in the first 18 weeks in previous years as a control group for non-covid-19 deaths in the 18 first weeks in 2020. The “treatment” period starts in week 10 of the year, when the first covid-19 death occurred in England and Wales.^15^ Summary statistics are presented in Table 1.

**Table 1.**
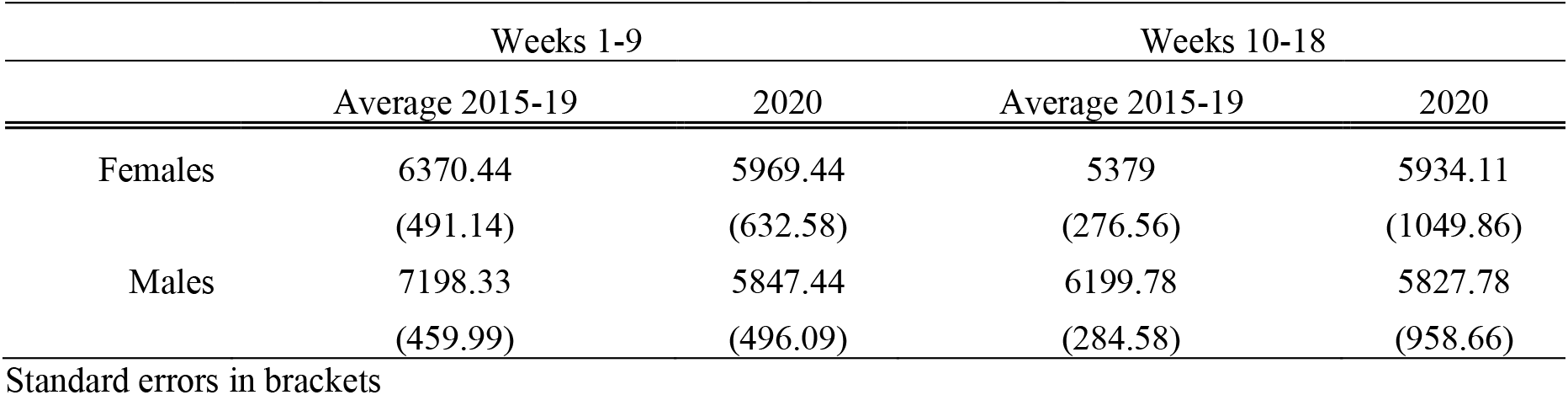
– Summary Statistics, number of weekly deaths not registered as covid-19-related

In order to compare trends in deaths *excluding covid-19 deaths* to the control group, I used a difference-in-differences (D-I-D) econometric approach. I used the average number of deaths in the previous five previous years as a control group, which also helps smooth out any short-term spikes (possibly due to a bad flu season^16^). A difference-in-differences approach requires that the trends (rather than absolute values) in treatment and control groups are parallel prior to the intervention. To test whether this common trend assumption is met, I followed the approach by Autor,^17^ who used a model with interactions including lags and leads (prior to and after the treatment). Results of the common trend assumption test are presented in Table A1 in the Appendix. All interaction *lags* are insignificant, suggesting that there is indeed a common trend in the two groups prior to the intervention. Trends can also be observed graphically (Figure 1).

**Figure 1.**
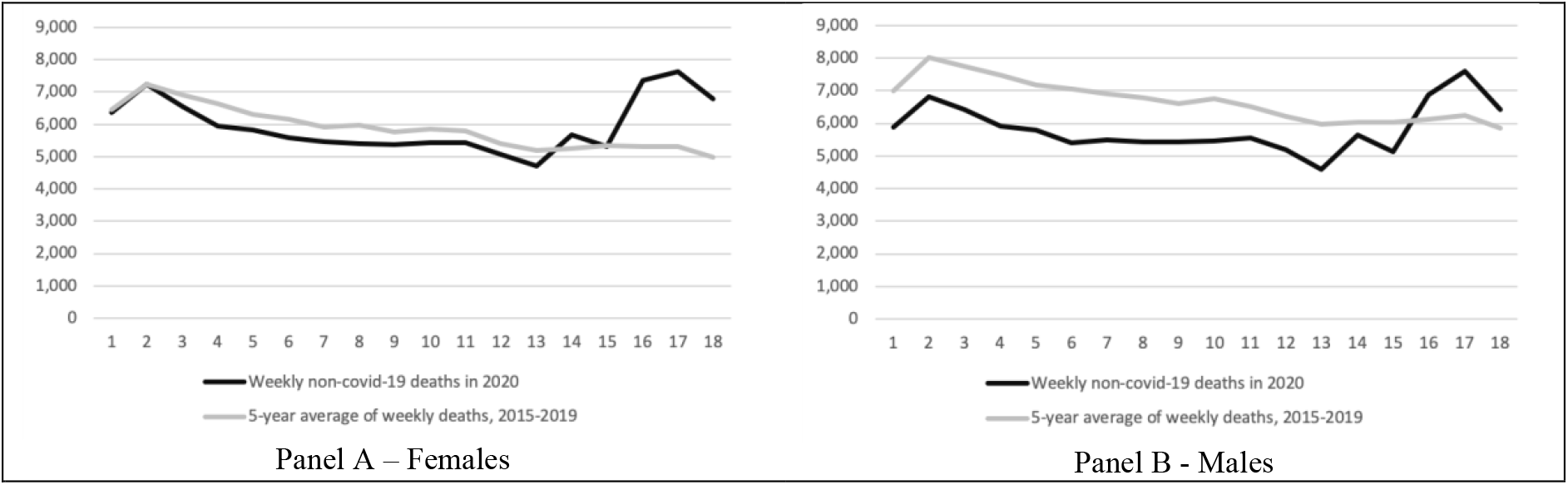
- Weekly deaths in England and Wales not registered as covid-19, first 18 weeks, year 2020 and average of years 2015-2019. First covid-19 death in week 10. Based on ONS data published on 12 May 2020.

The dependent variable is the number of deaths in each of the 18 first weeks of the calendar years, excluding any deaths that mentioned covid-19 in the death certificate. The difference-in-differences model includes a “treatment group” dummy variable, which takes the value of 1 for the group that is affected by the intervention, and zero otherwise. In this case, observations in 2020 take the value of 1, and observations in previous years take the value of zero. Another dummy that is included is an “after” variable, which takes the value of 1 in the period after an intervention (for both groups, 2020 and other years), and zero otherwise. We consider the treatment period to start in week 10, as that is the week when the first covid-19 death was reported, thus indicating an escalating situation and capturing any spillover effects of the virus. There were five covid-19-related deaths reported in week 10; 41 in week 11; 397 in week 12; 1,838 in week 13; 5,079 in week 14; 8,073 in week 15; 8,121 in week 16; 6,746 in week 17 and 4744 in week 18. One might argue that the treatment period should start later, when the number of deaths started increasing steeply, but a question that remains is where we should draw the line, and this would possibly relate with the cause of excess deaths, which is currently unknown – so identifying where the treatment period should start becomes particularly challenging. To be on the safe side, I followed the most conservative approach, i.e. a treatment period that starts with the first death, rather than when the number of deaths demonstrate large increases. This might underestimate the magnitude of any effect on non-covid-19 deaths, but is unlikely to exaggerate any findings.

The interaction of these two dummy variables (treatment*after) is the main variable of interest. I also used dummy variables for gender and week dummies, to address seasonality. Robust standard errors were used in all regressions.

Finally, for completeness, I also employed an interrupted time series model, using *all* weekly observations (not the average) from the first week of 2015 until the 18th week of 2020 (279 observations in total). Again, the treatment period starts in the tenth week of 2020, which does indeed leave a very short post-treatment period (9 weeks) compared to the pre-treatment period (270 weeks). However, this approach is used as an additional check rather than as the main analysis.

## 3. Results

There are 3,268 additional deaths that did not officially involve covid-19 in week 18 of 2020, compared to the same week in years 2015-2019 on average. Figure 1 shows the weekly number of deaths by gender in England and Wales (excluding any covid-19 deaths) in the first 18 weeks of 2020 and the average weekly deaths for the period 2015-2019. On week 14, 2020, onwards, there is a jump in non-covid-19 deaths, compared to the trend in previous years. This increase only started in week 14, i.e. in the fifth week of reported covid-19 fatalities. In week 13 2020 (which was the fourth week of covid-19 fatalities), the number of non-covid-19 deaths demonstrated a relative decrease. Figure A1 in the Appendix provides trends in non-covid-19 deaths by age group and gender, for age groups 65-74; 75-84; and 85 or over, which account for over 85% of all deaths.

Results of the baseline difference-in-differences econometric analysis are presented in Table 2, where weekly deaths enter the model by gender. There is an increase in deaths not reported as covid-19-related in the post-treatment period compared to the control group [D-I-D coeff: 967.50; 95%CI: 470.55 to 1464.45].

**Table 2.**
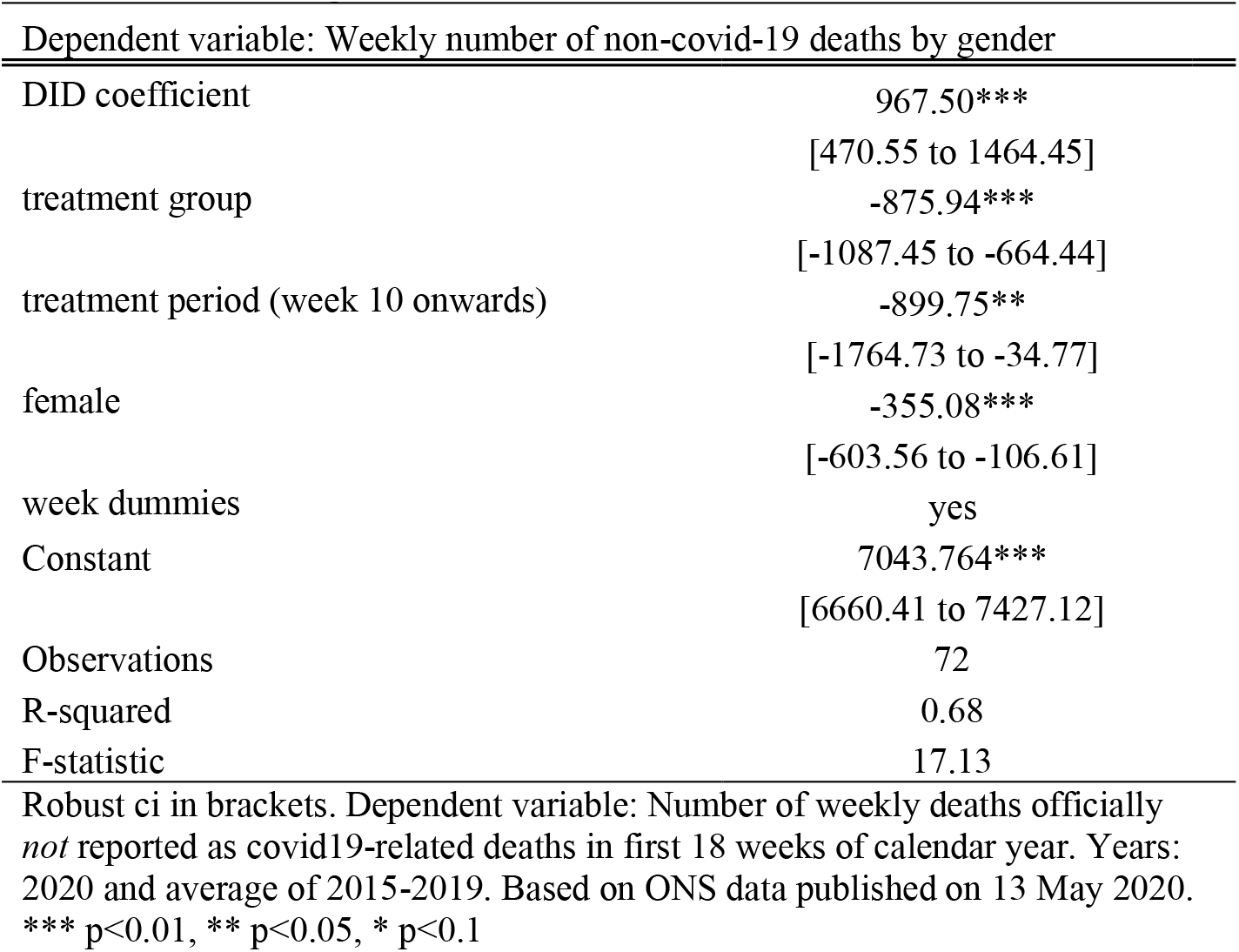
– D-i-D regression results

| Dependent variable: Weekly number of non-covid-19 deaths by gender |  |
| --- | --- |
| DID coefficient | 967.50***<br>[470.55 to 1464.45] |
| treatment group | -875.94***<br>[-1087.45 to -664.44] |
| treatment period (week 10 onwards) | -899.75**<br>[-1764.73 to -34.77] |
| female | -355.08***<br>[-603.56 to -106.61] |
| week dummies | yes |
| Constant | 7043.764***<br>[6660.41 to 7427.12] |
| Observations | 72 |
| R-squared | 0.68 |
| F-statistic | 17.13 |

Robust ci in brackets. Dependent variable: Number of weekly deaths officially *not* reported as covid19-related deaths in first 18 weeks of calendar year. Years: 2020 and average of 2015-2019. Based on ONS data published on 13 May 2020.
\*\*\* p<0.01, \*\* p<0.05, \* p<0.1

Could this relative increase in deaths be random? To answer this, I performed a placebo test, restricting the sample to the pre-treatment period (up to week 9, i.e. before the first covid-19 death), using an earlier random treatment period starting in week 7. Finding no effect in this case would lend additional support that the findings of the baseline model are not random. Results are provided in Table 3, and indeed, there is no effect in this placebo regression [D-I-D coeff: -23.08; 95%CI: -463.44 to 417.27].

**Table 3.** Placebo test

| Dependent variable: Weekly number of non-covid-19 deaths by gender |  |
| --- | --- |
| Placebo DID coefficient | -23.08<br>[-463.44 to 417.27] |
| treatment group | -868.25***<br>[-1161.50 to 575.00] |
| placebo treatment period | -625.21**<br>[-1144.79 to -105.63] |
| female | -352.94***<br>[-577.03 to -128.86] |
| week dummies | yes |
| Constant | 7038.85***<br>[6659.23 to 7418.46] |
| Observations | 36 |
| R-squared | 0.87 |
| F-statistic | 24.08 |

Robust CI in brackets. Dependent variable: Weekly number deaths not registered as covid-19-related by gender in first 9 weeks of calendar year. Years: 2020 and average of 2015-2019. Placebo treatment: Week 7 onwards. Based on ONS data published on 5 May 2020. \*\*\* p<0.01, \*\* p<0.05, \* p<0.1

I also performed the analysis at the Region level (Wales and nine regions in England). Results are reported in Table A2 in the Appendix, and confirm the findings of the baseline model [D-I-D coeff: 166.90; 95%CI: 122.06 to 211.74].

Results of the interrupted time series analysis are reported in Table A3 in the Appendix and suggest a similar effect, indicating an increase in the weekly deaths trend compared to the pre-treatment period [coef: 656.78; 95% CI: 410.63 to 902.94].

## 4. Discussion

This paper studied whether, during the covid-19 pandemic, there was an increase in deaths that have not officially been linked to the virus. Using a differences-in-differences econometric approach by comparing trends in 2020 to the average trends in the previous five years, I find that there are an additional 968 weekly deaths not officially registered as covid-19 compared to what would have been expected in the absence of the pandemic. Therefore, apart from the official covid-19 death toll, there are additional deaths that might be somehow linked, either directly or indirectly, to covid-19.

There are two possible reasons for this excess mortality. First, some people might have died from covid-19 without being diagnosed. Second, there may be spillover effects on other causes, such as patients postponing treatment for unrelated health conditions in order to avoid contracting the virus in hospitals or GP clinics; prioritisation of covid-19 patients by health services; stress and anxiety related to the current financial and public health environment; domestic violence; and lack of activity or other effects due to the lockdown.^2-10^ This relative increase in deaths occurs despite reasonably expecting a reduction in some types of mortality, such as motor vehicle collisions, crime, pollution and smoking.

The way deaths are reported or registered is central to this research question. In the ONS data, coronavirus deaths are those that mention covid-19 on the death certificate, meaning that one may have died due to other causes, after having tested positive for covid-19. In any case, reporting deaths is challenging, and misclassification can often play a role in empirical results.^18-20^ Furthermore, weekly data updates often include revisions on provisional figures from previous weeks.

It is worth noting that the increase in deaths seems to only occur from week 14 onwards, i.e. in the fifth week since the first covid-19 death in the country - when there were already over 2,000 covid-19 deaths in England and Wales. For any excess deaths that might have been a result of not visiting a hospital or GP clinic due to fear of contracting a disease, it may be that either people changed their behaviour only when covid-19 deaths started rising steeply, or that any untreated health issues led to death with a time lag. Using a treatment period in the empirical model that would start later into the pandemic would show an even larger magnitude of non-covid-19 deaths.

If people are dying of covid-19 without being diagnosed, mortality may actually be higher than what we currently think. If people without covid-19 are also dying as a result of the virus, we to need act urgently, to minimise these tragic spillover effects. Data access on the causes of non-covid-19 deaths would allow us to understand the mechanism behind this phenomenon and would help design appropriate targeted responses to save lives.

## Data Availability

Data availability: The data used in this study are freely available online from the Office for National Statistics.

## None Funding

None

## Ethics approval

The data used were aggregate anonymous data from a public source so ethics approval was not required.

## Data availability

The data used in this study are freely available online from the Office for National Statistics.

## Checklist

There is no relevant checklist of observational studies.

## APPENDIX

**Table A1.** Testing the common trend assumption: Time-varying regression. Years 2020 and average of 2015-2019

| Dependent variable: Weekly number of non-covid-19 deaths |  |
| --- | --- |
| female | -355.08***<br>[-524.10 - -186.06] |
| treatment group | -603.50<br>[-1,481.27 - 274.27] |
| week dummies | yes |
| *lags:* |  |
| year2020*week2 | 4.00<br>[-1,238.97 - 1,246.97] |
| year2020*week3 | -224.50<br>[-1,354.70 - 905.70] |
| year2020*week4 | -526.00<br>[-1,609.16 - 557.16] |
| year2020*week5 | -334.50<br>[-1,443.81 - 774.81] |
| year2020*week6 | -507.50<br>[-1,683.05 - 668.05] |
| year2020*week7 | -331.00<br>[-1,486.72 - 824.72] |
| year2020*week8 | -368.00<br>[-1,413.69 - 677.69] |
| year2020*week9 | -164.50<br>[-1,221.96 - 892.96] |
| *leads:* |  |
| year2020*week10 | -261.50<br>[-1,363.08 - 840.08] |
| year2020*week11 | -60.00<br>[-1,055.40 - 935.40] |
| year2020*week12 | -78.50<br>[-1,111.31 - 954.31] |
| year2020*week13 | -319.00<br>[-1,407.69 - 769.69] |
| year2020*week14 | 611.00<br>[-450.69 - 1,672.69] |
| year2020*week15 | 133.50<br>[-978.40 - 1,245.40] |
| year2020*week16 | 1,987.50***<br>[683.94 - 3,291.06] |
| year2020*week17 | 2,454.50***<br>[1,314.75 - 3,594.25] |
| year2020*week18 | 1788.00***<br>[518.99 - 3057.01] |
| Constant | 6,907.54***<br>[6,689.81 - 7,125.27] |
| Observations | 72 |
| R-squared | 0.90 |
| F-statistic | 13.16 |

Robust ci in brackets. Dependent variable: Number of weekly deaths *not* registered as covid-19-related in first 18 weeks of calendar year. Years: 2020 and average of 2015-2019. Based on ONS data published on 12 May 2020. \*\*\* p<0.01, \*\* p<0.05, \* p<0.1

**Table A2.** – D-i-D regression results by Region

| Dependent variable: Weekly number of non-covid-19 deaths by region |  |
| --- | --- |
| DID coefficient | 166.9***<br>[122.06 to 211.74] |
| treatment group | -45.23***<br>[-61.86 to -28.61] |
| treatment period (week 10 onwards) | -148.07***<br>[-223.81 to -72.34] |
| Region dummies | yes |
| week dummies | yes |
| Constant | 1351.17***<br>[1304.61 to 1397.73] |
| Observations | 360 |
| R-squared | 0.92 |
| F-statistic | 208.46 |

Robust ci in brackets. Dependent variable: Number of deaths per week not registered as covid19-related by region in first 18 weeks of calendar year. Years: 2020 and average of 2015-2019. Based on ONS data published on 12 May 2020. \*\*\* p<0.01, \*\* p<0.05, \* p<0.1

**Table A3.**
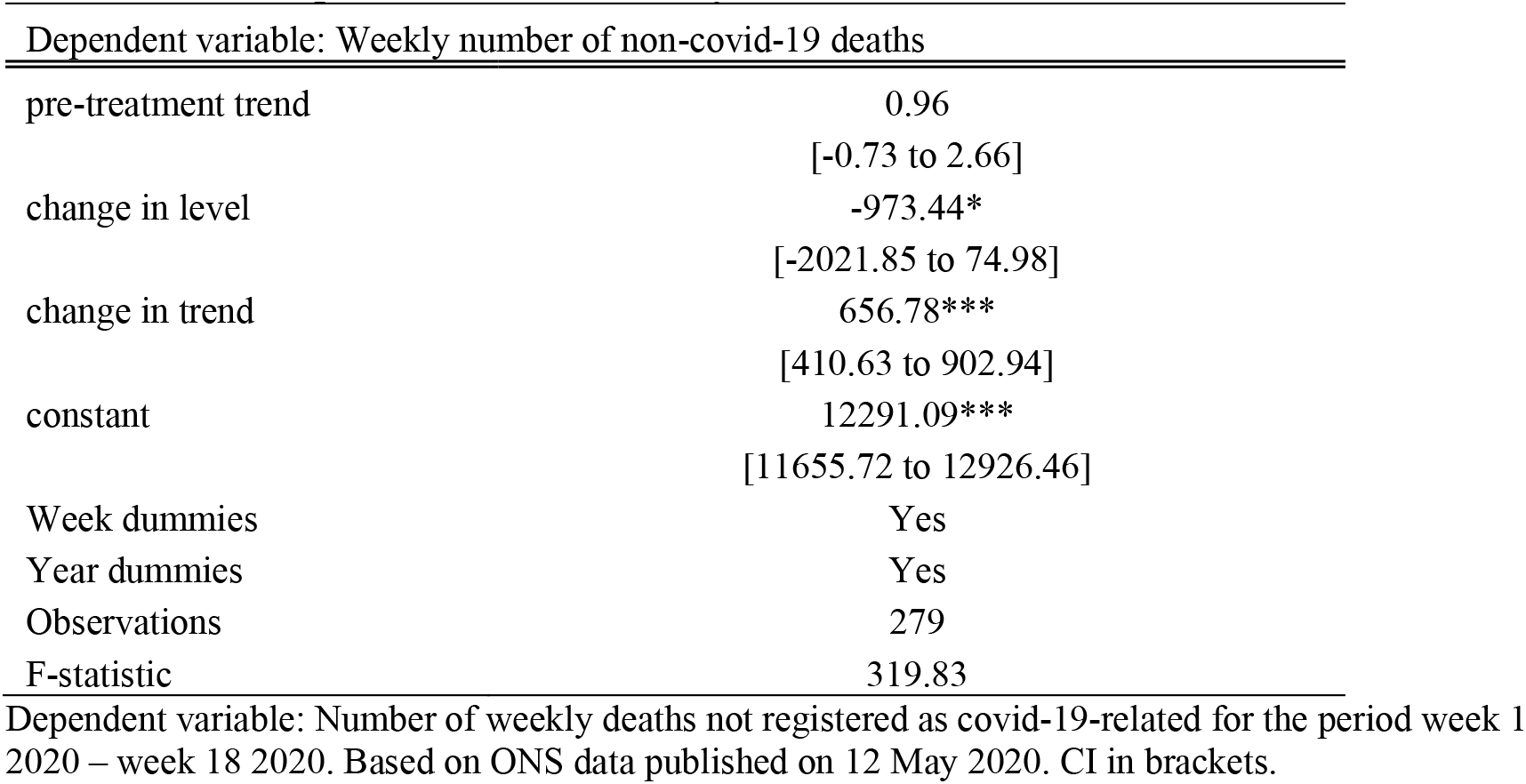
Interrupted Time Series Analysis

| Dependent variable: Weekly number of non-covid-19 deaths |  |
| --- | --- |
| pre-treatment trend | 0.96<br>[-0.73 to 2.66] |
| change in level | -973.44*<br>[-2021.85 to 74.98] |
| change in trend | 656.78***<br>[410.63 to 902.94] |
| constant | 12291.09***<br>[11655.72 to 12926.46] |
| Week dummies | Yes |
| Year dummies | Yes |
| Observations | 279 |
| F-statistic | 319.83 |

Dependent variable: Number of weekly deaths not registered as covid-19-related for the period week 1 2020 – week 18 2020. Based on ONS data published on 12 May 2020. CI in brackets.

**Figure A1.1.**
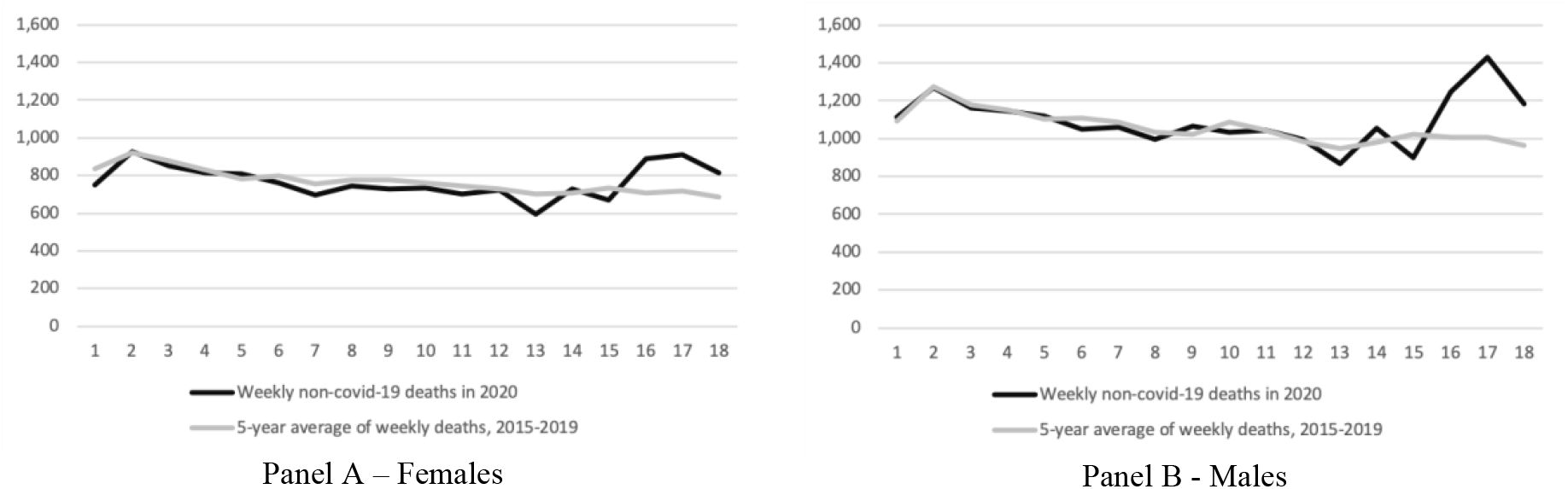
- Weekly deaths in England and Wales not registered as covid-19-related, first 18 weeks, year 2020 and average of years 2015-2019, age group 65-74

**Figure A1.2.**
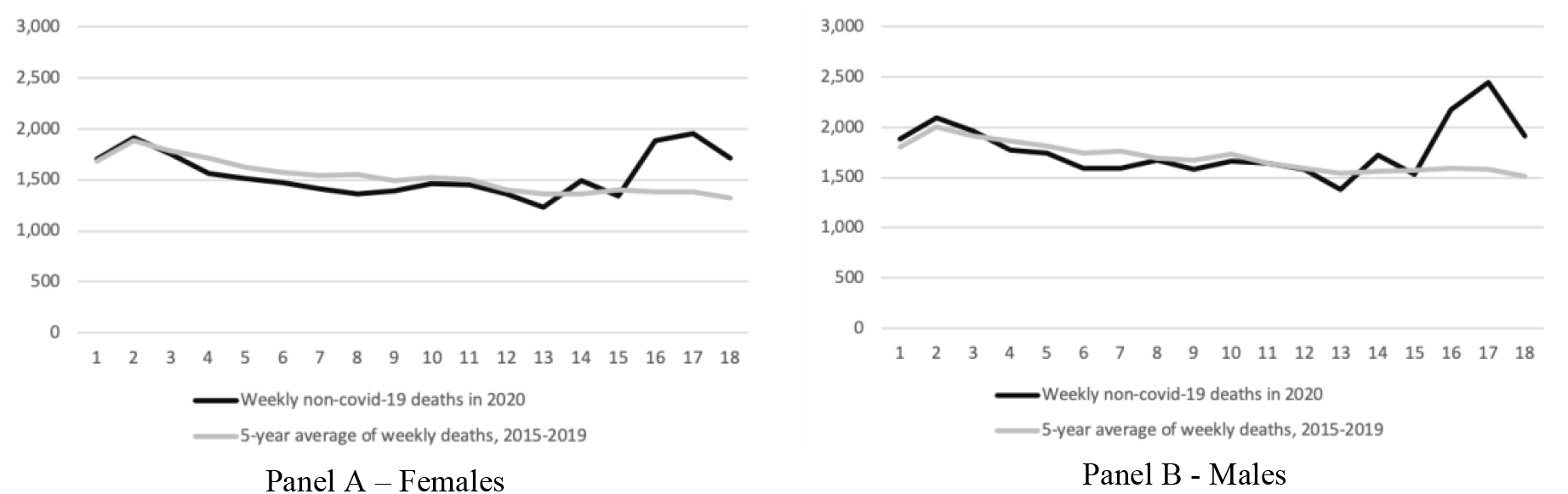
- Weekly deaths in England and Wales not registered as covid-19-related, first 18 weeks, year 2020 and average of years 2015-2019, age group 75-84

**Figure A1.3.**
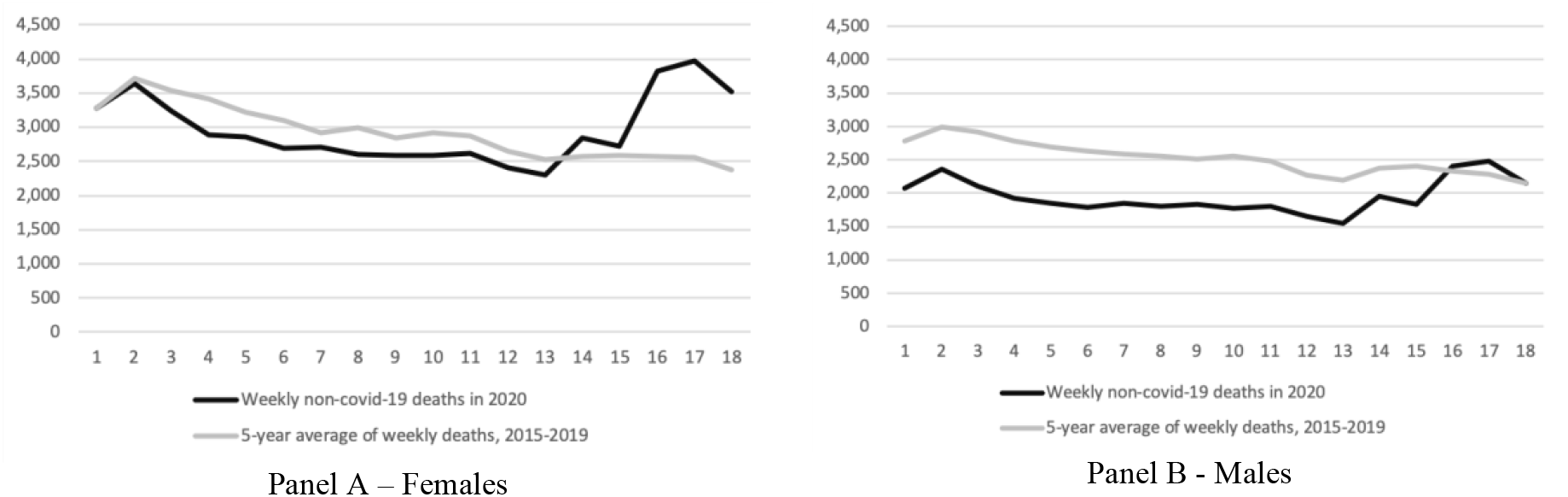
- Weekly deaths in England and Wales not registered as covid-19-related, first 18 weeks, year 2020 and average of years 2015-2019, age group 85+

## References

[1] Johns Hopkins University (2020). Covid-19 Dashboard by the Center for Systems Science and Engineering. https://coronavims.jhu.edu/map.html Accessed 22 April 2020.

[2] New York Times 2020a. Where have all the heart attacks gone? Available at: https://www.nytimes.com/2020/04/06/well/live/coronavirus-doctors-hospitals-emergency-care-heart-attack-stroke.html Accessed 14 April 2020.

[3] The Guardian. 2020. NHS to postpone millions of operations to tackle coronavirus. Available at: https://www.theguardian.com/society/2020/mar/17/nhs-postpone-millions-operations-tackle-coronavirus Accessed 14 April 2020.

[4] Kawachi, I. and Berkman, L.F., 2001. Social ties and mental health. Journal of Urban health, 78(3), pp.458–467.

[5] Metcalfe, R., Powdthavee, N. and Dolan, P., 2011. Destruction and distress: using a quasiexperiment to show the effects of the September 11 attacks on mental well-being in the United Kingdom. The Economic Journal, 121(550), pp.F81–F103.

[6] Lahti-Koski, M., Pietinen, P., Heliövaara, M. and Vartiainen, E., 2002. Associations of body mass index and obesity with physical activity, food choices, alcohol intake, and smoking in the 1982-1997 FINRISK Studies. The American journal of clinical nutrition, 75(5), pp.809–817.

[7] Saxena, S., Van Ommeren, M., Tang, K.C. and Armstrong, T.P., 2005. Mental health benefits of physical activity. Journal of Mental Health, 14(5), pp.445–451.

[8] New York Times, 2020b. A New Covid-19 Crisis: Domestic Abuse Rises Worldwide. Available at: https://www.nytimes.com/2020/04/06/world/coronavirus-domestic-violence.html April 6, 2020. Accessed 14 April 2020.

[9] Vandoros, S., Avendano, M. and Kawachi, I., 2019. The association between economic uncertainty and suicide in the short-run. Social Science & Medicine, 220, pp.403–410.

[10] Noelke, C. and Avendano, M., 2015. Who suffers during recessions? Economic downturns, job loss, and cardiovascular disease in older Americans. American journal of epidemiology, 182(10), pp.873–882.

[11] Di, Q., Wang, Y., Zanobetti, A., Wang, Y., Koutrakis, P., Choirat, C., Dominici, F. and Schwartz, J.D., 2017. Air pollution and mortality in the Medicare population. New England Journal of Medicine, 376(26), pp.2513–2522.

[12] Office for National Statistics. 2020. Deaths registered weekly in England and Wales, provisional. Available at: https://www.ons.gov.uk/peoplepopulationandcommunity/birthsdeathsandmarriages/deaths/datasets/weeklyprovisionalfiguresondeathsregisteredinenglandandwales Accessed 7 April 2020 and 14 April 2020.

[13] Department of Health and Social Care 2020. Number of Coronavirus cases and risk level in the UK. Available at: https://www.gov.uk/guidance/corona.virus-covid-19-information-for-the-public Accessed 14 April 2020.

[14] Powdthavee, N., Plagnol, A.C., Frijters, P. and Clark, A.E., 2019. Who got the Brexit blues? The effect of Brexit on subjective wellbeing in the UK. Economica, 86(343), pp.471–494.

[15] Coronavirus: Woman in 70s becomes first virus fatality in the UK https://www.bbc.com/news/uk-51759602 Retrieved 14 April 2020.

[16] National Flu Reports. Available at: https://www.gov.uk/government/statistics/weekly-national-flu-reports-2019-to-2020-season

[17] Autor, D.H., 2003. Outsourcing at will: The contribution of unjust dismissal doctrine to the growth of employment outsourcing. Journal of Labor Economics, 21(1), pp.1–42.

[18] Vandoros, S., 2020. The Association between Florida’s Opioid Crackdown and Opioid-Related Mortality: The Role of Economic Factors and Mortality Misclassification. American journal of epidemiology. DOI: doi org/10.1093/aje/kwaa016

[19] Flegal, K.M., Kit, B.K. and Graubard, B.I., 2018. Bias in hazard ratios arising from misclassification according to self-reported weight and height in observational studies of body mass index and mortality. American journal of epidemiology, 187(1), pp.125–134.

[20] Kapusta, N.D., Tran, U.S., Rockett, I.R., De Leo, D., Naylor, C.P., Niederkrotenthaler, T., Voracek, M., Etzersdorfer, E. and Sonneck, G., 2011. Declining autopsy rates and suicide misclassification: a cross-national analysis of 35 countries. Archives of general psychiatry, 68(10), pp.1050–1057.

